# A MATLAB algorithm to automatically estimate the QT interval and other ECG parameters and validation using a machine learning approach in congenital long-QT syndrome

**DOI:** 10.1101/2023.11.07.23298155

**Authors:** Elinor Tzvi-Minker, Sven Dittmann, Andreas Keck, Eric Schulze-Bahr

## Abstract

**Objective:** We present a novel MATLAB^®^ algorithm designed to automatically estimate the QT interval as well as other parameters in digital electrocardiograms (ECGs) of patients with congenital long QT syndrome (LQTS).

**Background:** Normal myocardial repolarization and the QT interval (i.e., the duration from the beginning of the QRS complex to the end of the T wave) are crucial markers for arrhythmogenesis, diagnosis and monitoring of LQTS. Manual measurement is time consuming and prone to physician’s variability and potential errors, necessitating an efficient and precise automated approach.

**Methods:** Our algorithm implements Lepeschkin’s tangent method to determine the T-wave end and, subsequently, the QT interval. Based on this, the algorithm additionally calculates other ECG parameters and T-wave characteristics: T-peak to T-end, Twave area, T-wave duration and R-peak to T-peak amplitude ratio. The results are two-fold validated: (1) against expert measurement of the QT interval (QT_GS), as well as (2) against the algorithm results of the ECG machine used (QT_MU). Machine-learning methods are applied to investigate the accuracy of the estimated parameters by our algorithm in a cohort of 363 LQT1 patients.

**Results:** Estimation of T-wave end, the QT interval and other ECG parameters by our algorithm was successful in at least one of the three ECG channels in 362 out of 363 patients (99.7%). On the other hand, the ECG machine-based QT_MU algorithm could estimate the QT interval only in 245/363 (95.3%) of recordings. Individual differences between the developed MATLAB^®^ algorithm in assessment of the QT and QT_GS were 10-fold more accurate to differences between QT_MU and QT_GS. Applying an optimizable Ensemble classifier on 30 ECG parameters extracted by our MATLAB^®^ algorithm had an accuracy of 74.91% and area under the curve (AUC) of 0.82 in classifying LQTS patients with a prolonged QTc interval (upon QT_GS) from those with a normal QTc interval.

**Conclusion:** The presented MATLAB^®^-based algorithm offers a robust and reliable approach to automatic QT interval estimation and QTc calculation in LQTS patients, potentially improving diagnostic precision and patient management. The results suggest that integrating further ECG parameters can assist with clinical assessment of LQTS patients. Future development will focus on integration into mainstream ECG devices and broadening the applicability to other arrhythmic conditions.

## Introduction

Long-QT Syndrome (LQTS) is a genetically heterogeneous, inherited cardiac arrhythmia syndrome, typically characterized by prolongation of the QTc interval on the surface electrocardiogram (ECG). In addition, T-wave abnormalities, and a propensity to develop polymorphic ventricular tachyarrhythmias are common. These may cause triggered syncope, sudden cardiac arrest, or death. In addition to congenital forms, also acquired conditions are known to cause secondary and potentially clinically relevant forms of disturbance in myocardial repolarization. From a clinical point-of-view, early detection of LQTS or abnormal myocardial repolarization is crucial to implement a tailored treatment, prevent life-threatening arrhythmias, detect genetic subforms and offer genetic counseling. The latter can reduce the risk of sudden cardiac death (SCD) in affected individuals and related, potentially affected, family members. Correct and precise detection of LQTS in the surface ECG can be, however, challenging and resemble a random finding, e.g., when associated with non-specific symptoms, in the setting of intermittent, heart-rate dependent ECG changes, including so-called concealed QTc prolongation (Doldi et al., 2022; Sugrue et al., 2016), and due to a genetic complexity (Ingles et al., 2020). Furthermore, the lack of routine ECG screening (Van Der Werf et al., 2010) and reliance on manual measurements or automatic estimations by ECG analysis software, leads to unrecognized or misjudged diagnoses of LQT syndrome. Of note, a correct determination of the QT interval is not homogeneous, even by clinical experts (Kashou et al., 2023).

The QT interval is defined between the onset of the Q-wave and the offset of the Twave in the surface ECG, measured in a baseline 12-lead or exercise ECG. The Bazett formula is commonly used to correct the measured QT interval by the heart-rate (HR) (so-called QTc interval duration), and is preferred over other corrections methods for LQT1 and LQT2 (Dahlberg et al., 2021), two genetic subtypes that are together causative for 30-35% of all LQTS cases (Adler et al., 2020). At higher HR, the Bazett’s formula tends to result in longer QTc values, whereas at lower HR, QTc values are shorter when compared to other methods of correction.

The T-wave end, which marks the end of ventricular repolarization, is a critical component in determining the QT interval. Thus, accurate and automated detection of the T-wave end is imperative for correct diagnosis and effective monitoring and management of LQTS patients. Traditionally, the determination of the T-wave end has been reliant on manual measurements, which are time-consuming and susceptible to intra- and inter-observer variability. Indeed, most physicians, including cardiologists, have difficulties to correctly identify a prolonged QT interval (Postema & Wilde, 2014), probably due to the complexity and variability of T-wave morphologies in LQTS patients and genetic subforms. To date, different methods have been used to correctly assess the T-wave end (Moeyersons et al., 2017; Vink et al., 2018). Importantly, Lepeschkin’s ‘tangent method’ has been shown to be unaffected by the electrical heart axis orientation and applied on a beat-to-beat basis regardless of the T-wave morphology. It is therefore the preferred method for T-wave end determination by numerous electrophysiologists (Vink et al., 2018).

Recently, several automated algorithms have been developed to detect the T-wave end, based on a variety of measures (Hermans et al., 2017). Of note, current commercial ECG management systems measure the QT interval by automated algorithms based on the average of a median complex over a period of time (Kligfield et al., 2018). Consequently, beat-to-beat detection algorithms which include the QT-interval dynamicity have been published, but often use only a single ECG lead (mostly II or V5), which makes the QT interval susceptible to heart axis orientation and electrode placement.

Here, we developed a MATLAB^®^ (2021a, The MathWorks, Natick, MA, USA)-based algorithm to specifically and automatically detect the end of the T-wave in three channels (II, V2 and V5) as a marker of repolarization in individual beats using the ‘tangent method’. Thereby, the MATLAB^®^ algorithm automatically determines various ECG parameters such as the QT interval, HR (for subsequent calculation of the QT interval corrected according to Bazett (QTc (B)) and corrected according to Fridericia (QTc (F)), T-peak to T-end, the T-wave duration, and the T-wave area. In addition, the R-peak to T-peak amplitude ratio is also calculated.

These unique parameters that reflect T-wave morphology may additionally contribute to the clinical expression of LQTS. Notably these are usually not estimated in commercial systems. We then investigate the contribution of these parameters to a correct evaluation of long-QT syndrome using MATLAB^®^ based machine-learning methods.

## Methods

### 1. Clinical data and ECG recordings

In this study, anonymized 12-lead ECG recordings in supine position from 363 LQTS patients were included. Patients had confirmed pathogenic or likely pathogenic variant in the *KCNQ1* gene (genetic LQT1 subtype). Data were collected during routine outpatient care at the Institute for Genetics of Heart Diseases of the University Hospital Münster. The study was conducted in accordance with the Declaration of Helsinki (as revised in 2013) and approved by institutional ethics board of University Hospital Münster, Münster, Germany (2021-315-f-S). An informed consent was signed by all study participants.

### 2. Data acquisition and pre-processing

Pre-processing steps and subsequent analyses of the raw ECG data are shown in Fig. 1. 12-lead ECGs were recorded for 10 sec with a 500 Hz sampling frequency using an ECG machine (GE CAM-HD Registrator, GE Cardiosoft V6.73, GE HealthCare). The data was stored using the MUSE™ data management system (Muse v9 Cardiology Information System, GE HealthCare) and later exported in an anonymized form in XML format. Subsequently, XML files were converted into CSV files using freely available Python script^1^.

**Fig. 1.**
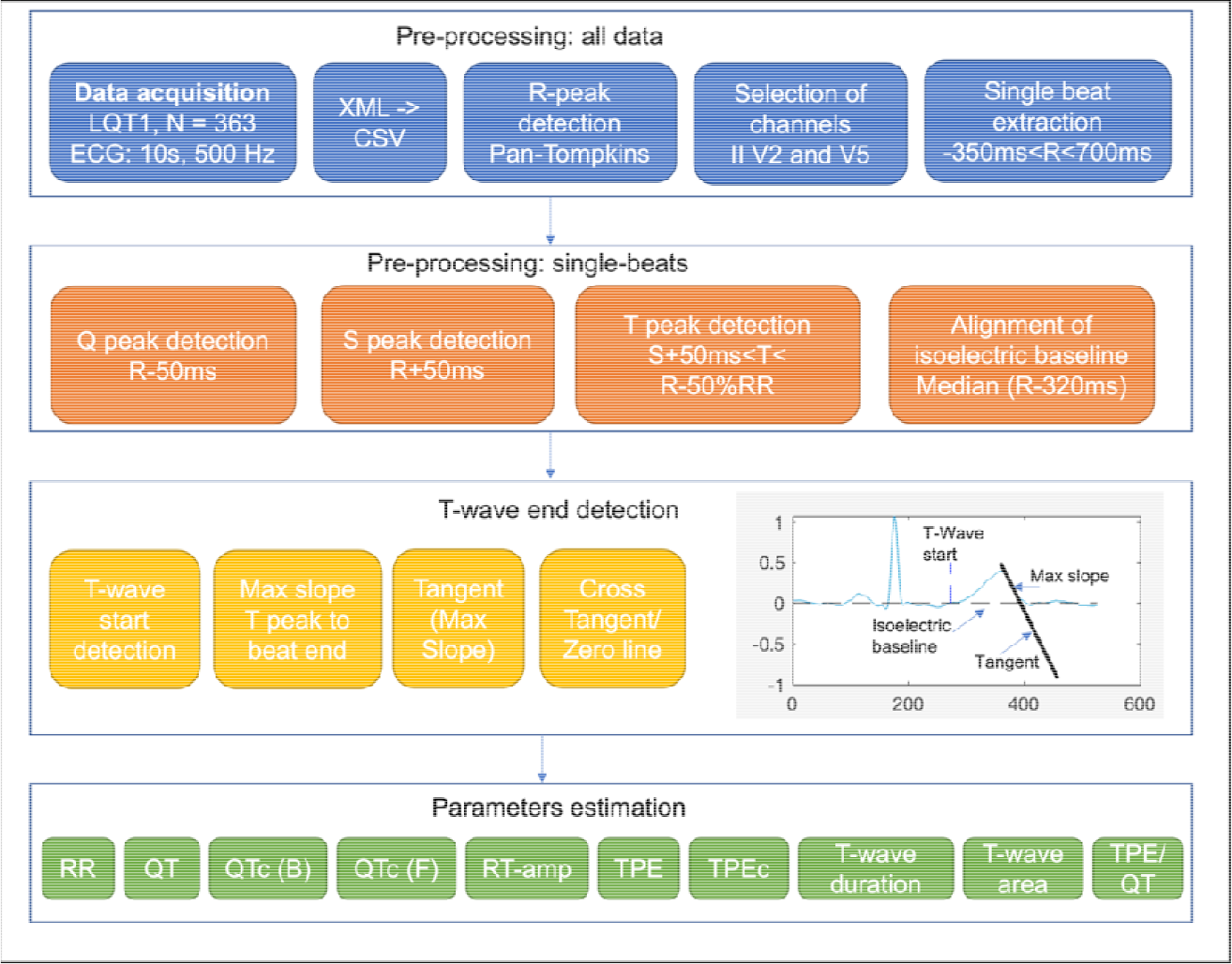
Overview of the pre-processing and analysis steps for automated detection of the t-wave end and evaluation of ECG parameters.

**Fig. 2.**
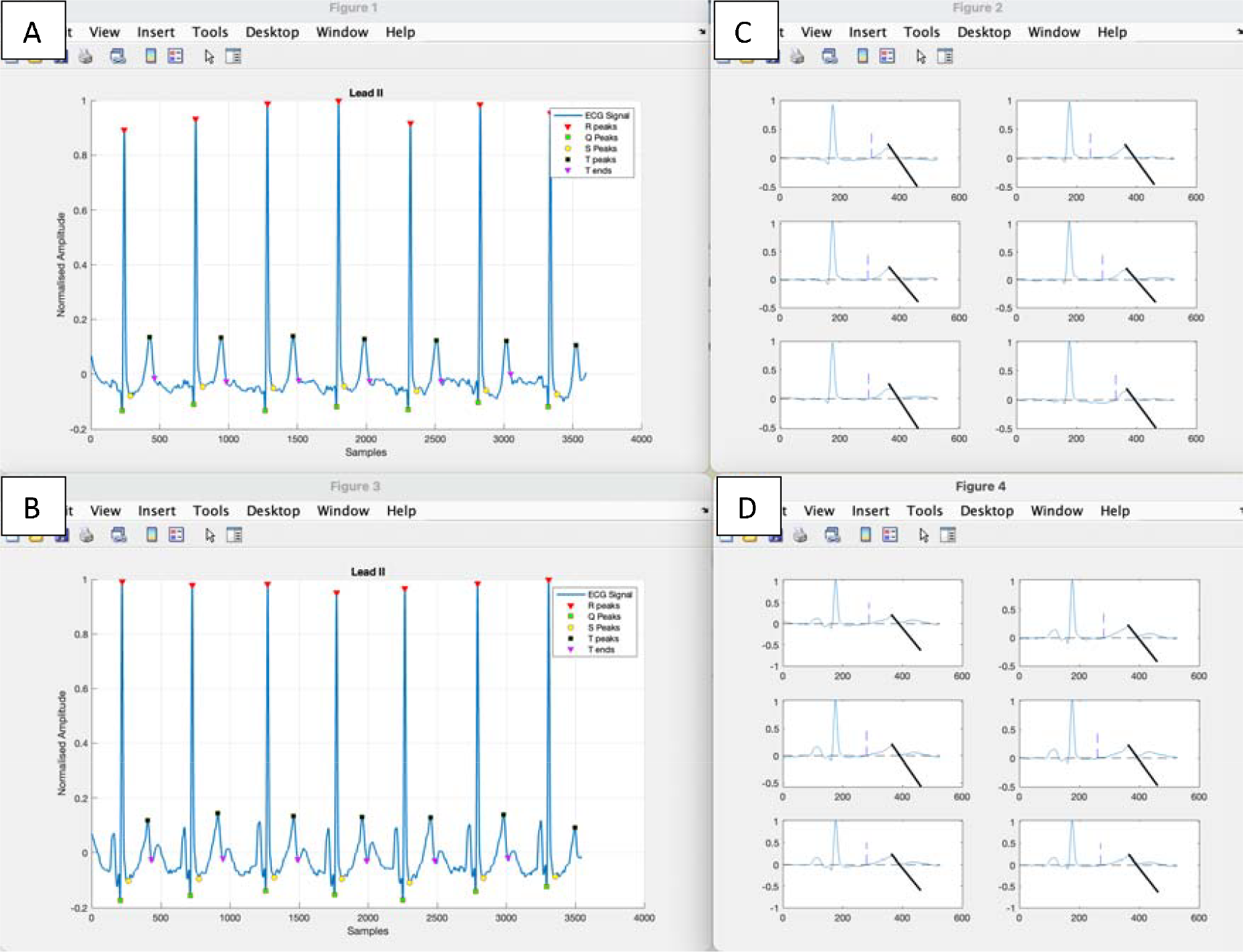
Plots produced by the MATLAB^*®*^ algorithm in two exemplary patients (Patient 1: A,C; Patient 2: B, D). Plots A-B show the normalized ECG from Lead II with the respective annotations for QRS peak locations (green, red and yellow resp.) as well as T start and T end locations (black and purple resp.). Plots C-D show the beat-by-beat detection of the T-wave end according to the tangent method (the tangent is annotated as a black line).

### 3. Description of the algorithm

Next, a method to detect accurately the T-wave end was developed using custommade MATLAB^®^ scripts and open-source routines.

(https://de.mathworks.com/matlabcentral/fileexchange/66098-ecg-p-qrs-t-wave-detecting-matlab-code).

The following pre-processing steps were taken:

1. R-peaks were identified using a MATLAB^®^ implementation of Pan-Tompkins algorithm^2^.
2. Beats were windowed according to the R-peak locations with 350 ms before R-peak until 700 ms after the R-peak.

Next, the QRS as well as other ECG markers were searched for three lead channels II, V2 and V5:

1. Q- and S-peaks were defined as the local minimum within a 50 ms window before and after the R-peak respectively.
2. T-peak was searched within a delay of 50 ms following S-peak and 50% of the RR distance (in sec) before the next R-peak. We accounted for both, negative and positive peaks.
3. For detection of the T-wave start and T-wave end, we followed these steps:
  1. Isoelectric baseline of single beats was aligned with the zero line by subtracting the median of the first 320 ms of each beat.
  2. T-wave start was defined to be the first point within a search window between S-wave peak and T-wave peak that crossed the isoelectric line (adjusted to whether the T-wave peak is positive or negative).
  3. The steepest point of the descending or ascending limb of the T-wave (depending on whether the peak is positive or negative) was searched using the differential of the amplitude within a time window between the T-wave peak and the end of the beat (defined above).
  4. The tangent through this point was calculated and the cross point between this line and the zero baseline was searched.
  5. The first sample of the signal following this point was marked as the Twave end.

### 4. Calculation of further ECG parameters

1. T-wave area was calculated using an approximation of the integral of the signal from T-wave start to end using the trapezoidal method (spacing: 2 ms). Note that the amplitude of the T-wave is not in voltages and is measured on a normalized ECG for the sake of cross-subject comparisons.
2. T-peak to T-end duration (TPE) was defined as the time between the T-wave peak and the T-wave end. The TPE was then HR-corrected as follows: 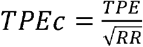=, with RR interval is 60/HR, calculated in seconds. We removed all data points that exceeded TPEc = 40 ms.
3. QT interval was defined as the duration between the beginning of the Q-wave peak (if absent: of the R-wave) and the T-wave end.
4. We then calculated the median of these values across all interval beats. The QTc interval was HR-corrected according to Bazett formula (QTc(B)): 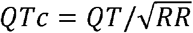 as well as according to the Fridericia formula (QTc(F)): 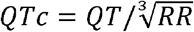 Whereas RR interval is 60/HR, calculated in seconds.
5. Next, we calculated the amplitude ratio between the R-peak and the T-peak.
6. Finally, the ratio TPE/QT (uncorrected) was calculated.

### 5. Validation of the algorithm

To assess the validity of our algorithm we extracted both the QT and QTc (corrected using the Fridericia method, i.e., QTc(F)) from the XML files of each patient, as assessed by the MUSE™ data management system. In addition, a manual measurement of the QT interval duration was independently performed by expert cardiologists to serve as a Golden Standard (GS) for comparisons with the algorithm output. We then calculated the differences in each patient between the QT as assessed by our algorithm (QT_a_) in channels II, V2 and V5 and QT assessed by the experts (QT_GS) to assess the validity of our algorithm. Similarly, we calculated the difference between QT as assessed by our algorithm (QT_a_) and QT as assessed by MUSE™ data management system (QT_MU).

Next, we assessed the probability of the algorithm to correctly detect prolonged QTc interval durations as diagnostic feature of LQTS using supervised machine-learning methods. Using machine-learning, we could assess the contribution of additional parameters (see above) beside the QTc in identification of normal QT or prolonged QTc values.

The genotype positive LQT1 patients were tagged according to their gender (female, male) and GS_QT values into two groups: those with a normal QTc(B), i.e., females: < 460 ms, males: < 450 ms, or prolonged QTc, i.e., females: > 460 ms, males > 450 ms.

As features we used the above mentioned 10 parameters: TPE, TPEc(B), TPE/QT, QT, QTc(B), QTc(F), RR interval, R-peak to T-peak amplitude ratio, T-wave area, and T-wave duration. Parameters were obtained from the three ECG channels II, V2 and V5, leading to a total of 30 parameters. We then used the classifier learner toolbox by MATLAB^®^ to investigate to which degree the ECG parameters can correctly distinguish LQT1 patients with normal QTc from those with a prolonged QTc. Subsequently, we tested this hypothesis with the following machine-learning algorithms: Support Vector Machine (SVM), k-nearest neighbors (KNN), Linear Discriminant Analysis (LDA), Decision Tree (DT), Logistic Regression (LR), and Naive Bayes (NB). The hyperparameters were set to optimizable when possible.

## Results

The manual assessment of T-wave end and subsequently the QT interval was performed in all LQTS patients (see Table 1). N – number of patients; SD - standard deviation; QT MU – assessment of QT by the MUSE™ algorithm, uncorrected. QT GS – Golden standard of QT manual measurement by experts, uncorrected. QTc (B) GS – Golden standard of QT manual measurement by experts, corrected using Bazett, QTc (F) GS – Golden standard of QT manual measurement by experts, corrected using Fridericia. QTc MU – assessment of QT by the MUSE™ algorithm, corrected using Fridericia.

**Table 1:**
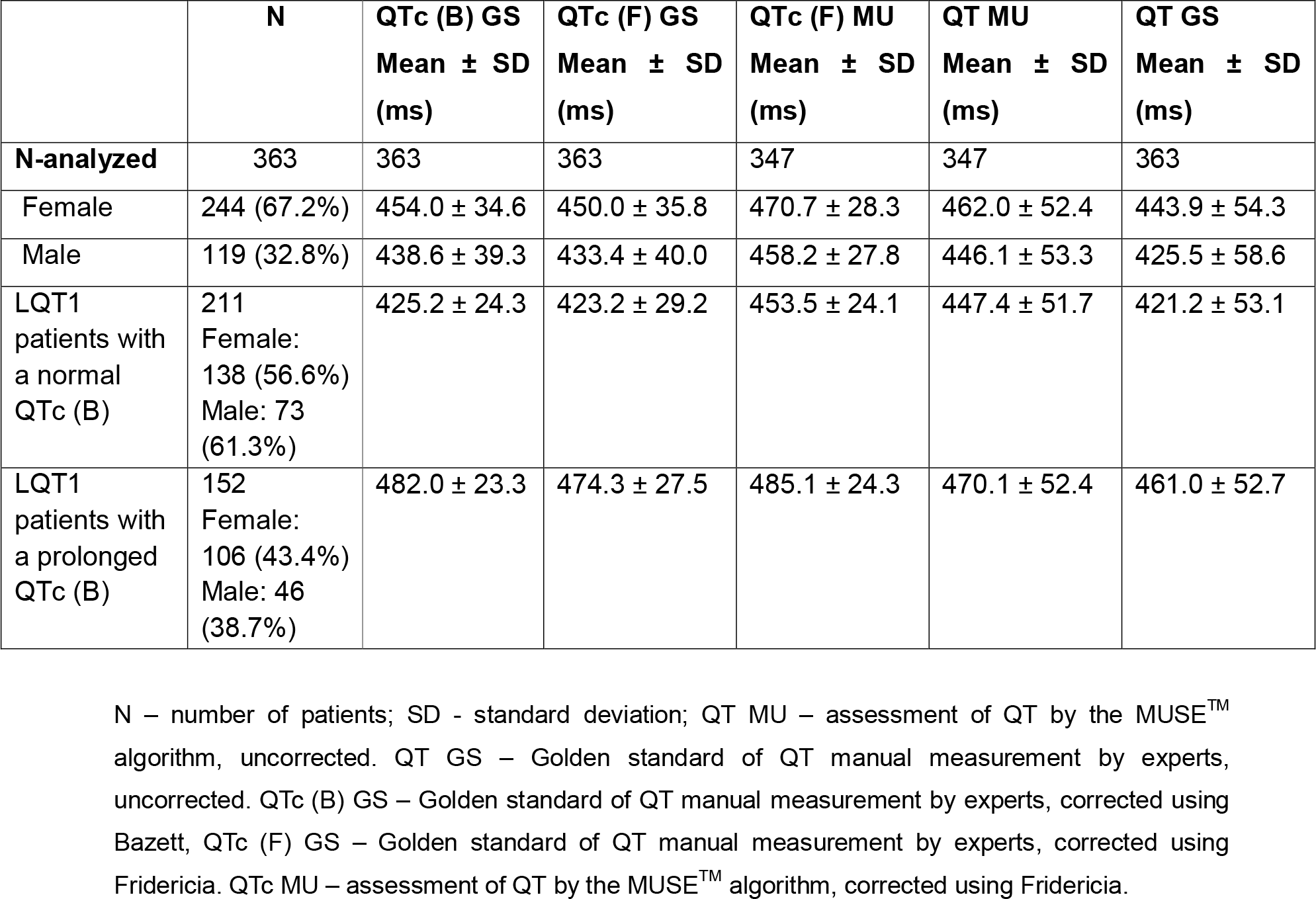
Demographics, manual and MUSE™ assessment of QTc values in LQT1 patients

**Table 2:** ECG parameters as assessed and calculated by MATLAB^®^ algorithm (mean ± SD) in 363 LQT1 patients

|  | ECG lead II | ECG lead V2 | ECG lead V5 |
| --- | --- | --- | --- |
| Total: N = 363 | 339 | 351 | 340 |
| QT<sub>a</sub> (ms) | 431.2 ± 62.6 | 437.3 ± 64.5 | 428.8 ± 61.4 |
| RR interval (ms) | 962.3 ± 214.4 | 960.6 ± 210.5 | 964.7 ± 211.6 |
| **QT<sub>a</sub>c (B) (ms)** | 442.3 ± 42.5 | 448.7 ± 46.2 | 439.5 ± 39.7 |
| **QT<sub>a</sub>c (F) (ms)** | 437.9 ± 44.7 | 444.3 ± 48.2 | 435.2 ± 42.7 |
| **TPE (ms)** | 69.2 ± 18.8 | 83.2 ± 24.4 | 69.0 ± 15.7 |
| **TPE to QT ratio** | 0.16 ± 0.03 | 0.19 ± 0.05 | 0.16 ± 0.03 |
| **TPEc (B) (ms)** | 73.3 ± 14.7 | 86.5 ± 23.4 | 72.7 ± 14.8 |
| **T-wave duration (ms)** | 240.4 ± 76.9 | 302.9 ± 77.6 | 260.2 ± 79.0 |
| **T-wave area (no unit)** | 11.8 ± 9.6 | 40.3 ± 23.0 | 9.5 ± 7.1 |
| **R to T amplitude ratio** | 2.48 ± 13.8 | 1.49 ± 1.9 | 3.42 ± 7.5 |

### MATLAB^®^ algorithm performance

The algorithm was able to determine the T-wave end and the other determined parameters in at least one of the three ECG channels across 362 out of 363 patients (99.7%); in the remaining patient, the ECG signal was too noisy to detect the T-wave peak. When assessing each ECG channel individually, the algorithm was able to assess the T-wave end in most measurements, i.e., for channel II in 341/362 patients (93.9%), for channel V2 in 352/363 patients (97.0%) and for channel V5 in 341/363 patients (93.9%).

The QT_MU interval was assessed in only 347/363 (95.6%) of the LQT1 patients. Thus, the MATLAB^®^ algorithm shows superiority in providing the QT interval in comparison to MUSE management system’s algorithm, since here no values have been provided in 16 LQT1 patients. We presume that the algorithm used by the MUSE™ management system to estimate the QT interval is based on one ECG channel estimation, which then might be inferior to the MATLAB^®^ algorithm that is based on three ECG channels. QT_a_ – QT interval without correction, QT_**a**_c (B) – QT interval corrected according to Bazett, QT_**a**_c (F) - QT interval corrected according to Fridericia, TPE – T-peak to T-end interval, TPEc (B) – TPE corrected according to Bazett.

To assess the performance of our algorithm in correctly estimating the QT interval, we calculated the individual differences between QT_a_ and QT_GS (uncorrected) per channel:

II: -7.6 ± 2.1 ms; V2: -1.8 ± 2.4 ms; V5: -9.6 ± 1.9 ms; (mean ± STD of the mean across patients). The difference between QT_MU and QT_GS across patients was: 16.9 ± 1.5 ms. Similarly, the individual differences between QT_a_C (F) and QTc_GS (uncorrected) was per channel: II: -6.2 ± 2.1 ms; V2: -0.8 ± 2.3 ms; V5: -8.7 ± 1.9 ms; (mean ± STD of the mean across patients). The difference between QTc (F)_MU and QTc (F)_GS across patients was: 16.6 ± 1.4 ms.

The distribution of single-subject differences to QT_GS are shown in the swarm plots in Figure 3. Similarly, we show in Figure 4 the single-subject differences to QTc (F)_GS. These results suggests that our algorithm assessment is a factor of 10 more accurate than that of MUSE™ management system. QT_MU will generally produce a longer QT compared to QT_GS, which in principle means an over-diagnosis of patients with long QT.

**Figure 3.**
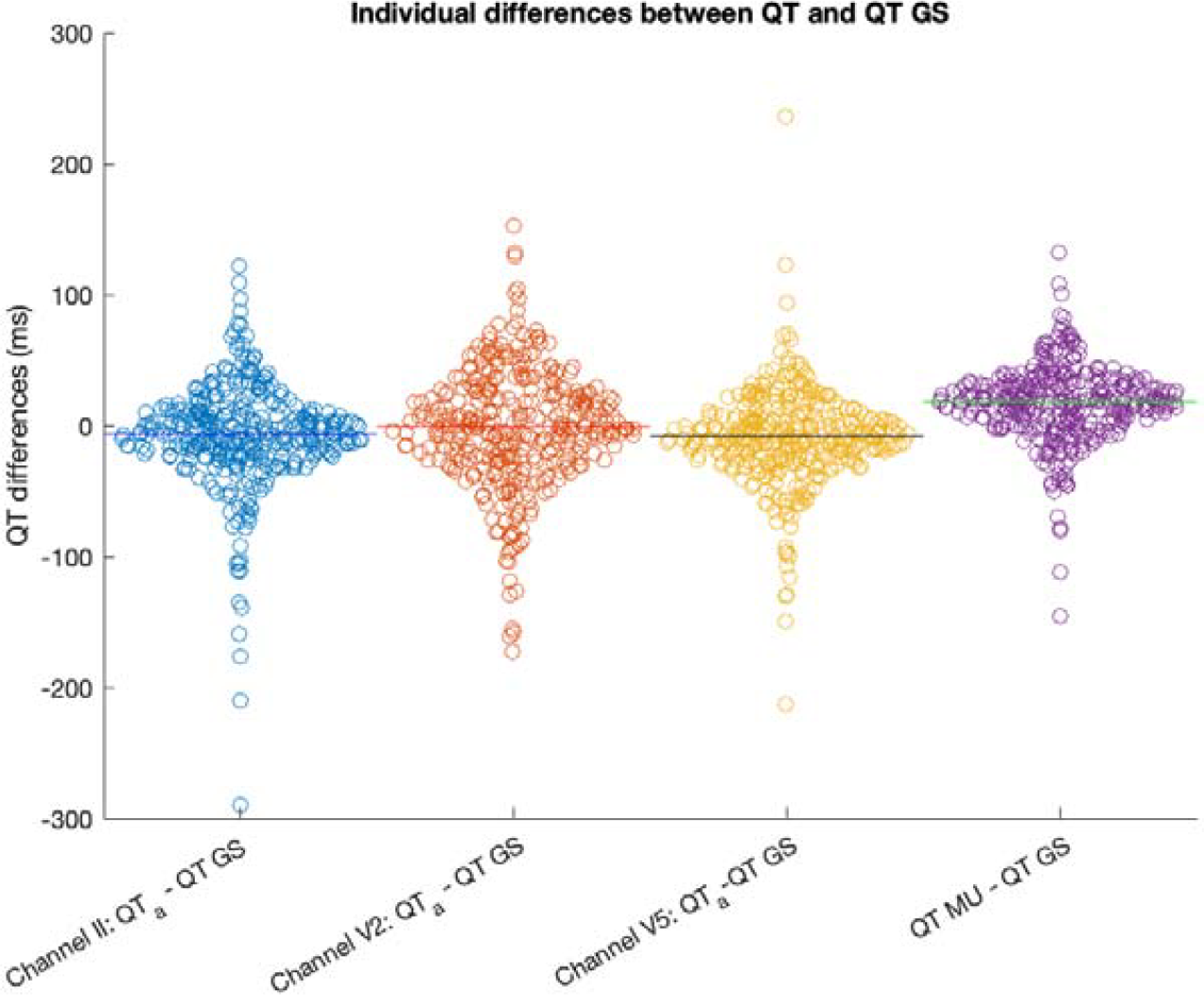
Violin plots show individual differences in QT interval estimation (uncorrected) of our MATLAB® algorithm (QT_a) in ECG channels II, V2 and V5 to expert manual estimation of QT interval (QT_GS uncorrected). For comparison individual differences between QT as assessed by MUSE™ management system (QT_MU uncorrected) and QT_GS (uncorrected) is displayed.

**Figure 4.**
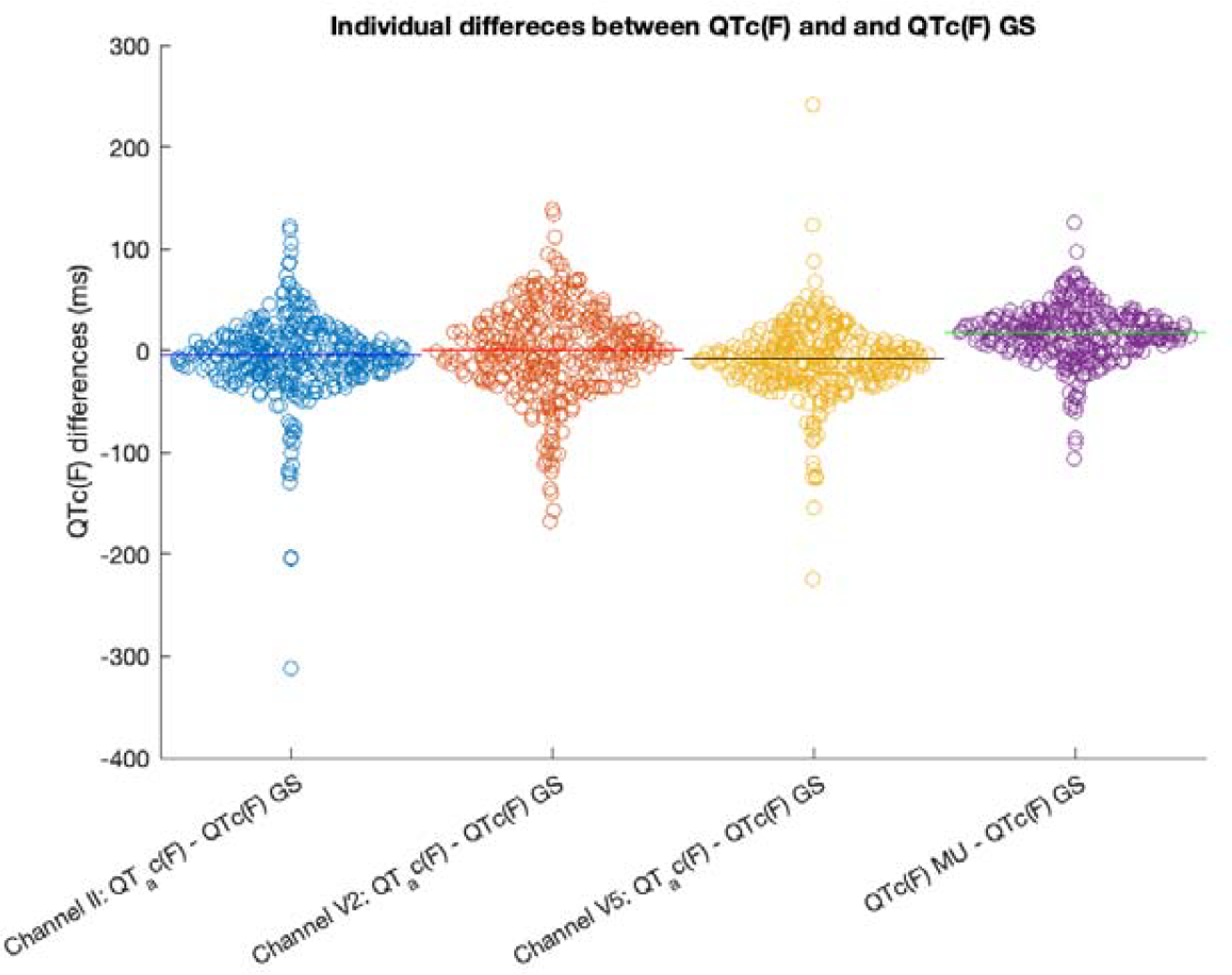
Violin plots show individual differences in QTc (corrected according to Fridericia) interval estimation of our MATLAB® algorithm (QT_a_c(F)) in ECG channels II, V2 and V5 to expert manual estimation of QTc interval (QTc(F)_GS). For comparison, individual differences between QTc as assessed by MUSE management system (QTc(F)_MU) and QTc(F)_GS is displayed.

### Validation of the algorithm using concealed LQT

When classifying all 30 ECG values (10 parameters x 3 channels), the best performing algorithm was an optimizable Ensemble that had an accuracy of 74.1% and AUC of 0.82 (see Figure 5 below). An Ensemble classifier combines a set of trained weak learner models and responds for new data by aggregating predictions from its weak learners. Bayesian optimization was performed with 30 iterations. The results of all classification algorithms are presented in Table 3.

**Table 3.** Classification accuracy and area under the curve (AUC) results for each tested algorithm

|  | Accuracy | AUC |
| --- | --- | --- |
| Optimizable decision tree (DT) | 71.6% | 0.74 |
| Optimizable linear discriminant analysis (LDA) | 71.6% | 0.79 |
| Logistic regression (LR) | 69.1% | 0.77 |
| Optimizable Naive Bayes (NB) | 73.3% | 0.81 |
| Optimizable Support vector machine (SVM) | 72.5% | 0.80 |
| Optimizable k-nearest neighbors (KNN) | 66.7% | 0.71 |
| **Optimizable Ensemble** | **74.1%** | **0.82** |

**Figure 5.**
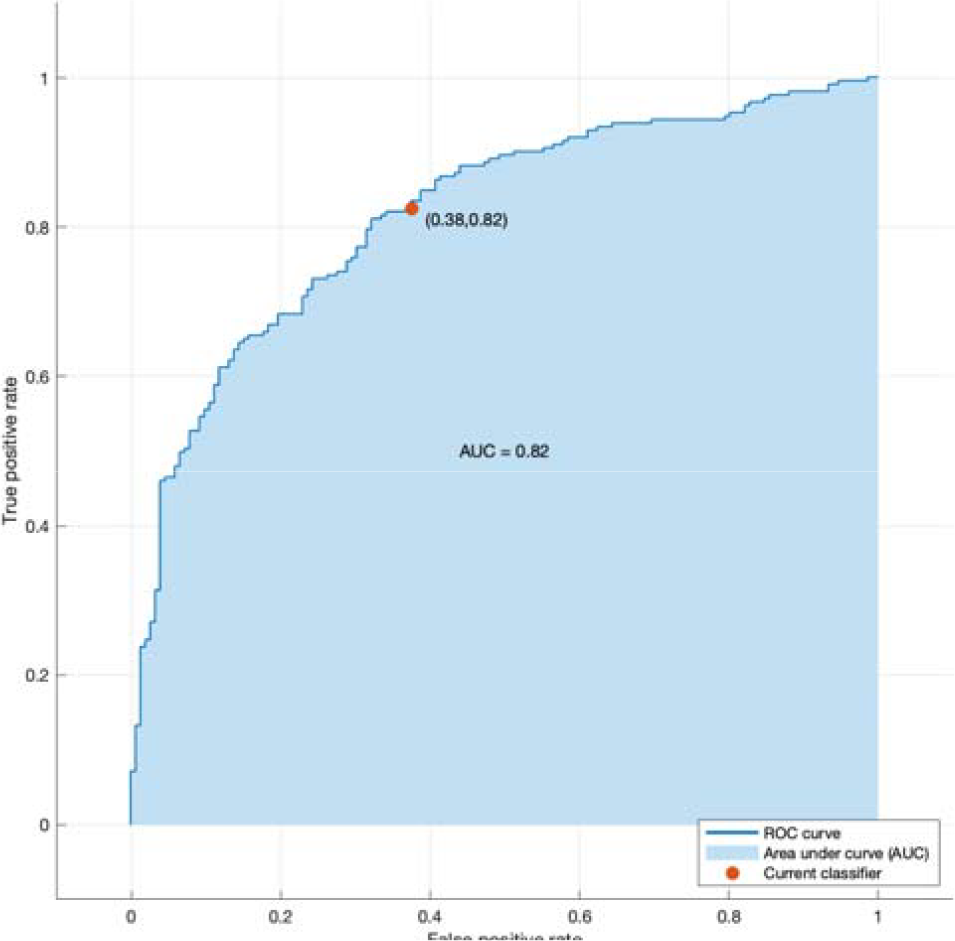
The receiver operating characteristic (ROC) curve showing true positive rate versus false positive rate for the trained Ensemble classifier.

Applying principal component analysis (PCA) to the winning model only worsens the results. Only 4 components were kept after training, resulting in 70.5% accuracy and AUC of 0.75. We also examined the model performance when only including the QTc (B) or the QTc (F) from the three channels. This has not improved the classification results (QTc(B): 73.8%, 0.80 AUC / QTc(F): 73.0%, 0.75 AUC).

## Discussion

The present study introduces a novel MATLAB^®^ algorithm that first automatically detects the T-wave end, and then subsequently estimates QT interval (corrected and uncorrected), T-peak to T-end (TPE, corrected and uncorrected), T-wave area, Twave duration and R-peak to T-peak amplitude ratio in digital ECGs. The performance of the algorithm was evaluated against manual measurements by specialists (as golden standard - GS) and the estimated QT by MUSE™ management system (QT_MU), in 363 LQT1 patients. Not only that our algorithm could assess the QT interval in more LQTS patients compared to the MUSE™ data management system, but most importantly the results demonstrate remarkable accuracy, being 10-fold more precise than QT_MU (both corrected and uncorrected). This exceptional performance has the potential to substantially reduce false positives, thus holding significant implications for clinical practice and patient care. In addition, we show that the use of morphological parameters of the T-wave as listed above additionally contribute to the correct assessment of LQTS and can immensely contribute to the clinical assessment and management of these patients.

Most modern ECG machines offer automated measurements of ECG intervals to users. However, implemented algorithms have some limitations. For instance, the QT-interval is determined based on an averaged complex over time, resulting in the loss of temporal fluctuations and the dynamic nature of the QT-interval concerning changes in heart rate (HR). Additionally, the lack of transparency regarding the specifics of these algorithms poses a challenge to their users. Nevertheless, despite this limitation, many cardiologists rely on and trust the QTc-interval provided by ECG machines. Here we show that relying on ECG machine output only could lead to misdiagnosis of patients with LQTS. The GE MUSE™ data management system, although widely used in clinical settings, relies on proprietary algorithms, and may suffer from limitations due to the nature of its design and the complexity of ECG signal analysis. Our algorithm’s improved performance signifies its potential to surpass existing commercial solutions and set a new benchmark for QT measurement as well as calculation of other ECG parameters.

Manual assessment of the QT interval is time consuming for a physician’s daily routine. Therefore, in practice, only one lead and one ECG complex often are selected for measurement. This is a questionable practice as one lead might not be representative. Indeed, results from this study clearly show a strong variability across channels, in terms of T-wave end detection. In general, automatic algorithms are objective, observer independent, and time sparing methods to overcome issues of manual measurements. Unfortunately, current implementations are either unavailable to the public or are not accurate enough. For example, (Hermans et al., 2017) developed an algorithm to evaluate the QT interval based on the automated tangent method that can be applied beat-by-beat similarly to our algorithm. The authors found a small bias of -1.88 ms to +3.39 ms when compared to manual measurement similarly to the results achieved here.

The importance of correct ECG assessment in LQTS has been demonstrated in studies that aimed to detect LQTS patients with concealed phenotype, which means that patients have a genetic mutation but present with otherwise normal QT interval. In a study by Bos et al., deep neural networks of 12-lead ECG were used to distinguish patients with LQTS from those who were evaluated for LQTS but discharged as healthy (Bos et al., 2021). The authors found that using QTc alone, AUC was 0.82, whereas using all 12-leads, 10 s each, as an input to a deep neural network algorithm led to an AUC of 0.9, suggesting that additional parameters in the ECG signal could help to better identify LQTS patients. This result has been replicated (Doldi et al., 2022) who showed using a novel convolutional neural network model resulting in a high accuracy rate (91.8%) in identification of concealed LQTS. In another study, (Aufiero et al., 2022) used neural networks on ECG data to show that genotype positive LQTS patients can be accurately identified based on the ECG data only. Note that unlike our study, in which we take specific ECG parameters and investigate their influence on correct LQTS detection, the use of neural networks does not allow to deduct which parameters exactly influenced the correct diagnosis of LQTS patients. Thus, conclusions regarding the specific mechanisms associated with ECG parameters are very limited.

### Clinical Implications

Congenital LQTS is a life-threatening condition that requires early detection and intervention to prevent sudden cardiac events. Our algorithm offers a potential tool for highly accurate early identification of patients at risk, allowing for timely clinical intervention and improved patient outcomes.

### Future Directions

While the proposed algorithm has demonstrated significant advancements in T-wave end detection, there are several avenues for further improvement and exploration. Increasing the dataset size and diversity could enhance the algorithm’s robustness and generalizability across various patient populations and ECG acquisition settings. The high accuracy of QT interval measurement is of utmost importance for diagnosis and risk assessment in cardiac patients not only for congenital LQTS but also for evaluating acquired LQTS, such as risk for life-threatening drug-induced torsade de pointes arrhythmias. The algorithm’s accuracy can lead to more targeted interventions, minimizing unnecessary or life-threatening treatments and interventions, thereby optimizing patient care and potentially reducing healthcare costs.

## Conclusion

In conclusion, the presented MATLAB^®^ algorithm for automatic T-wave end detection and estimation of QT interval as well as other relevant parameters in digital ECGs exhibits superior accuracy compared to the GE MUSE™ data management algorithm. Its potential to reduce false positives and identify congenital LQTS underscores its clinical significance in improving patient care and decision-making. This algorithm opens-up new possibilities for enhancing cardiac diagnostics and monitoring, and it holds great promise for future advancements in combined genetic and ECG analysis.

## Data Availability

All data produced in the present study are available upon reasonable request to the corresponding author.

## Acknowledgements

The algorithm was developed as part of project LQTS-NEXT, funded by European Joint Program on Rare Diseases (EJP RD). Study sponsors had no influence on any aspect of the study (experimental design, data collection, manuscript preparation, submission for publication). We thank all participating LQTS patients for their support and the supplied informed consent. Data management has been provided by Virginia Böhm, Nilüfer Tuncer, and Corinna Rickert.

## Author contributions

*ETM:* Data curation, Methodology, Software, Validation, Formel analysis, Writing-original draft, Visualization. *SD:* Resources, Data curation, Writing - review and editing, Project administration. *AK:* Conceptualization, Project administration, Writing-review and editing, Supervision, Funding Acquisition. *ESB:* Conceptualization, Methodology, Writing-review and editing, Supervision, Funding Acquisition.

## Footnotes

1 https://github.com/DFNOsorio/GEMuseXMLReader

2 https://www.researchgate.net/publication/313673153_Matlab_Implementation_of_Pan_Tompkins_ECG_QRS_detector

## Notes

### Competing Interest Statement

The authors have declared no competing interest.

### Author Declarations

Institutional ethics board of University Hospital Muenster, Muenster, Germany has approved this study

